# Comparison of semaglutide and lifestyle counseling for weight loss using electronic health records

**DOI:** 10.64898/2025.12.01.25341393

**Authors:** William Powell, Diego R Mazzotti, Stephen D Herrmann, Ann M Davis, Bettina Mittendorfer, Jared Bruce, Xing Song, Lemuel R Waitman

## Abstract

**Importance:** Although it is well-established that GLP-1 receptor agonists are highly effective in causing marked weight loss in randomized controlled trials, there is a need to evaluate their effectiveness in real-world settings and compare it with the outcomes of traditional lifestyle counseling.

**Objective:** To evaluate the effectiveness of semaglutide compared with lifestyle counseling alone for weight loss in diabetic and non-diabetic patients with obesity.

**Design:** A multi-site, retrospective, observational study using data from electronic health records in a time-to-event analysis for comparative effectiveness.

**Settings:** Data were collected from 13 health systems across the central United States part of the Greater Plains Collaborative (GPC).

**Participants:** Adults with a body mass index greater than or equal to 30 kg/m^2^ and no prior history of malignant cancer, pregnancy, or bariatric surgery receiving semaglutide or lifestyle counseling alone.

**Exposures:** Participants received either lifestyle counseling alone with at least three sessions within the first 16 weeks or were prescribed semaglutide for a minimum of 16 weeks.

**Main Outcome and Measures:** The primary outcome was incidence of 10% or greater reduction in weight from baseline.

**Results:** There were 3,927 eligible participants, with 615 taking semaglutide and 3,312 receiving lifestyle counseling. Following propensity score matching, each group (diabetic and non-diabetic) consisted of 120 and 495 participants respectively. Hazard ratios for 10% weight loss were significantly higher from semaglutide than for lifestyle counseling for non-diabetic (hazard ratio, 1.63 [95% CI: 1.32, 2.01; p<0.001]) and diabetic (hazard ratio, 2.45 [95% CI: 1.47, 4.09; p<0.001]) populations.

**Conclusion and Relevance:** The data from our study suggest that adjunct semaglutide therapy compared with lifestyle counseling alone is more effective in achieving at least 10% weight loss. This information may be used to inform clinical practice guidelines and stimulate future research on the long-term weight loss effectiveness of this class of drugs.

**Key Points:** *Question:* Does semaglutide, a novel glucagon-like peptide-1 receptor agonist, improve weight loss outcomes compared to lifestyle counseling alone in patients with obesity in a real-world setting?

*Findings:* After adjusting for possible confounders, patients treated with semaglutide had a greater incidence and speed of achieving 10% weight loss compared to those receiving lifestyle counseling alone.

*Meaning:* Compared with lifestyle counseling alone, semaglutide is a more effective weight loss option for patients with obesity. Future studies should determine the cost-benefit ratio, broader health benefits, and long-term weight loss maintenance of semaglutide therapy compared to or combined with lifestyle counseling.

## Introduction

About two-thirds of the adult population in the United States is living with obesity (defined as body mass index, BMI, equal to or greater than 30.0 kg/m^2^) and the prevalence continues to rise.^1^ Obesity is associated with multiple health problems, including cardiovascular disease, stroke, and diabetes.^2^ As such, management of obesity and weight loss for individuals with obesity remains a high priority in an increasingly obese population with a high cost of disease^3^. Lifestyle counseling consisting of diet and exercise is the cornerstone of weight managment^4^ and often results in only modest amounts (less than 5%) of weight loss.^5^ Novel pharmacological therapies, such as GLP-1 receptor agonist class drugs, that are highly effective in achieving marked (greater than 10%) weight loss in randomized controlled trials have high potential for utility in the field of weight management. However, the effectiveness of these drugs in real-world clinical settings is unknown.

Approved in 2017 for type 2 diabetes management^6^ and in 2021 for weight loss,^7^ the GLP-1 receptor agonist class drug semaglutide has seen widespread use. We evaluated the effects of lifestyle counseling alone compared with semaglutide therapy on body weight by conducting a multi-site, retrospective study using data from electronic health records in a time-to-event analysis for comparative effectiveness. Semaglutide is marketed under three brand names that are different forms of the same compound. Rybelsus® is a pill indicated to treat type 2 diabetes, Ozempic® is an injection indicated to treat type 2 diabetes, and Wegovy® is an injection intended to treat weight loss in people with obesity (with or without diabetes).Our study focused on patients prescribed Wegovy® only and used real-world electronic health record data from health systems throughout the Central United States that are part of the Greater Plains Collaborative (GPC). Evidence from large-scale real-life data on comparative effectiveness may help shape future obesity management clinical practice.

## Methods

### Study Design

The present study was a multi-site retrospective study using Electronic Health Records from 13 health systems across the central United States part of the Greater Plains Collaborative (GPC),^8^ a PCORnet^9^ Clinical Research Network. Data were obtained from the Greater Plains Collaborative Reusable Observable Unified Study Environment (GROUSE)^10^ environment, a unique data repository combining health records and claims data from GPC sites. Data were stored in PCORnet Common Data Model (CDM) format.^11^

### Inclusion Criteria

Participants meeting the predefined inclusion criteria were included in the study. For individuals receiving semaglutide, a minimum exposure of 16 weeks was required to accommodate dose escalation. Similarly, participants undergoing lifestyle counseling alone needed to have at least 3 distinct appointments in the first 16 weeks to ensure baseline adherence to the therapy sessions. All participants were adults over 18 years of age, and needed to have at least 6 months of prior clinical activity with their health system to establish baseline interaction. All participants required a baseline BMI of ≥ 30 kg/m^2^, with height measurements available in the electronic health records to facilitate this calculation. These inclusion criteria were designed to encompass individuals engaging in both therapeutic modalities with a BMI indicative of obesity, demonstrating their intent to pursue weight loss through the initiation of either semaglutide or lifestyle counseling alone.

### Exclusion Criteria

Individuals with a history bariatric surgery, pregnancy in the prior year, or malignant cancer in the prior year were excluded from the study. The exclusion timeframe spanned from one year prior to semaglutide or lifestyle counseling initiation to the first prescription or counseling session, aiming to capture an active diagnosis period. Participants were exclusively exposed to either semaglutide or nutrition counseling, without any overlap between the groups.

### Weight Tracking

Alongside the baseline exclusion criteria, individuals were monitored for weight measurements relative to their treatment exposure. To be eligible, participants needed to have a baseline weight measurement within 30 days before starting semaglutide or lifestyle counseling alone, as well as at least one measurement taken more than 16 weeks after initiation that could be utilized for weight change analysis. Data cleaning procedures were applied to the weight measurements dataset. Weight data were cleaned by removing unrealistic raw weight values, such as those below 13 pounds or exceeding 1000 pounds, were excluded from the analysis. Additionally, weight measurements showing a discrepancy of more than 10 pounds on the same day were also excluded. Furthermore, measurements indicating an implausible weight change over a short duration, such as changes exceeding 30 pounds within 30 days, were omitted. These measures were implemented to eliminate erroneous weight entries or those recorded in incorrect units (e.g., kilograms instead of pounds) in the electronic health record (EHR). Participants with eligible and plausible weight measurements were included for further analysis.

### Type 2 Diabetes Stratification

The study was stratified into two groups based on participants’ incidence of type 2 diabetes mellitus. Type 2 diabetes mellitus was calculated using an extension of the method derived from Wiese et. al^12^ using PCORnet data. Patients were classified as having type 2 diabetes if they (1) had an International Classification of Diseases (ICD)-9 or ICD-10 diagnosis of type 2 diabetes and a prescription of an anti-diabetic medication within 90 days following diagnosis, (2) an ICD-9 or ICD-10 diagnosis of type 2 diabetes and a hemoglobin A1C (HbA1c) value greater than or equal to 6.5% using LOINC codes within 90 days of diagnosis date, or (3) any antidiabetic prescription within 90 days of an elevated HbA1c value. Patients that did not have either a diabetic diagnosis, prescription, or elevated HbA1c were classified as non-diabetic and those who had only 1 out of 3 of those conditions were excluded due to indeterminate diabetes status.

### Baseline Covariates

In addition to weight tracking and exposure, relevant baseline covariates that could be used to identify and adjust for different potential confounding effects were obtained from the EHR. These covariates were organized into 3 categories: demographic, clinical, and health service utilization variables. Demographic variables included sex, race, and age. Clinical variables included initial BMI, hemoglobin A1C (HbA1c), as well as diagnoses and prescription information. Diagnoses included conditions relevant to weight loss and cardiometabolic health. A complete list is available in Table 1. Medications were grouped using RxNorm^13^ codes into relevant Cardiovascular, Psychiatric, Pain, and ISx/NSAID categories. Health Service Utilization variables included the number of ambulatory or acute care visits each patient had with their health system.

**Table 1:**
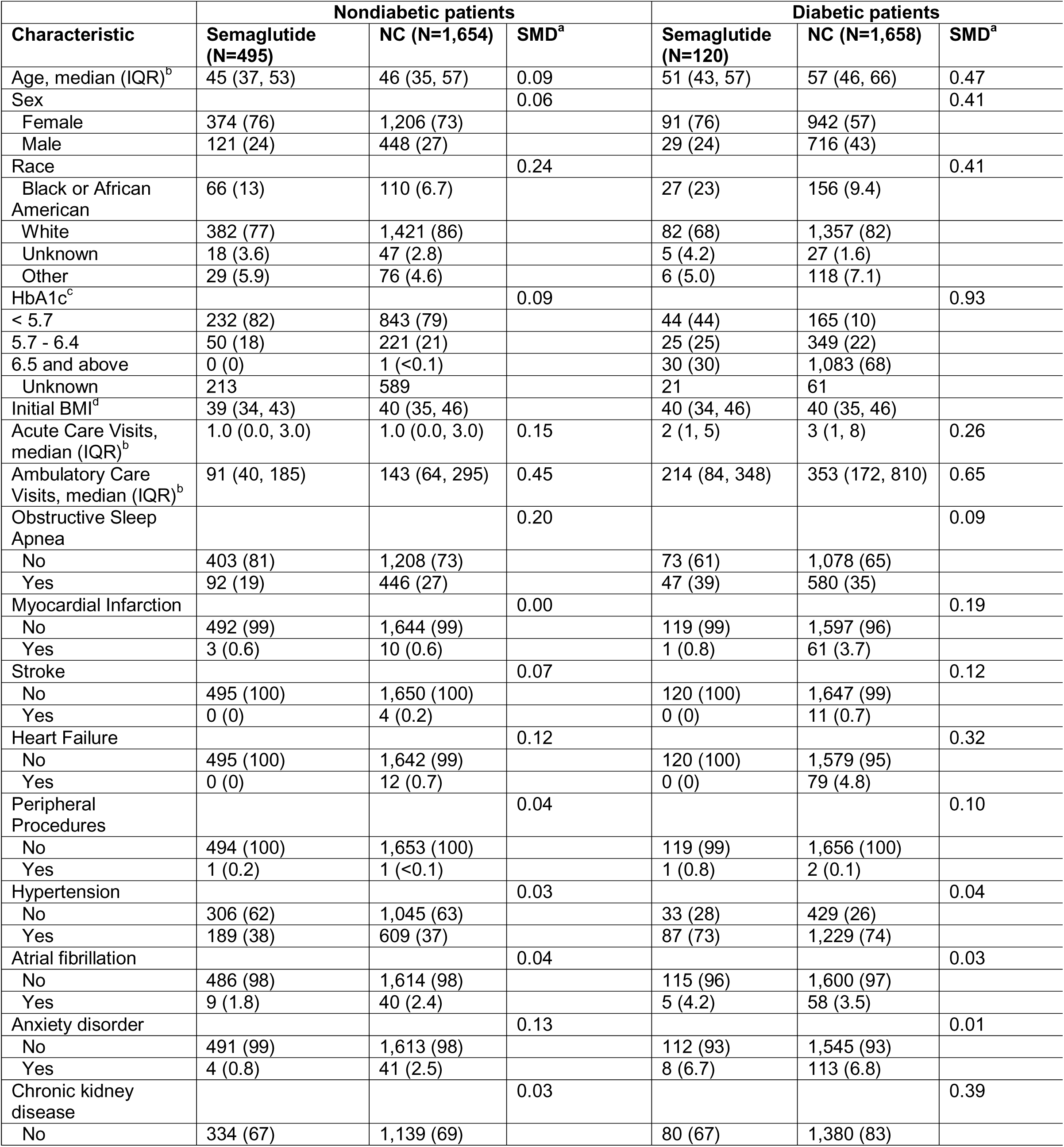
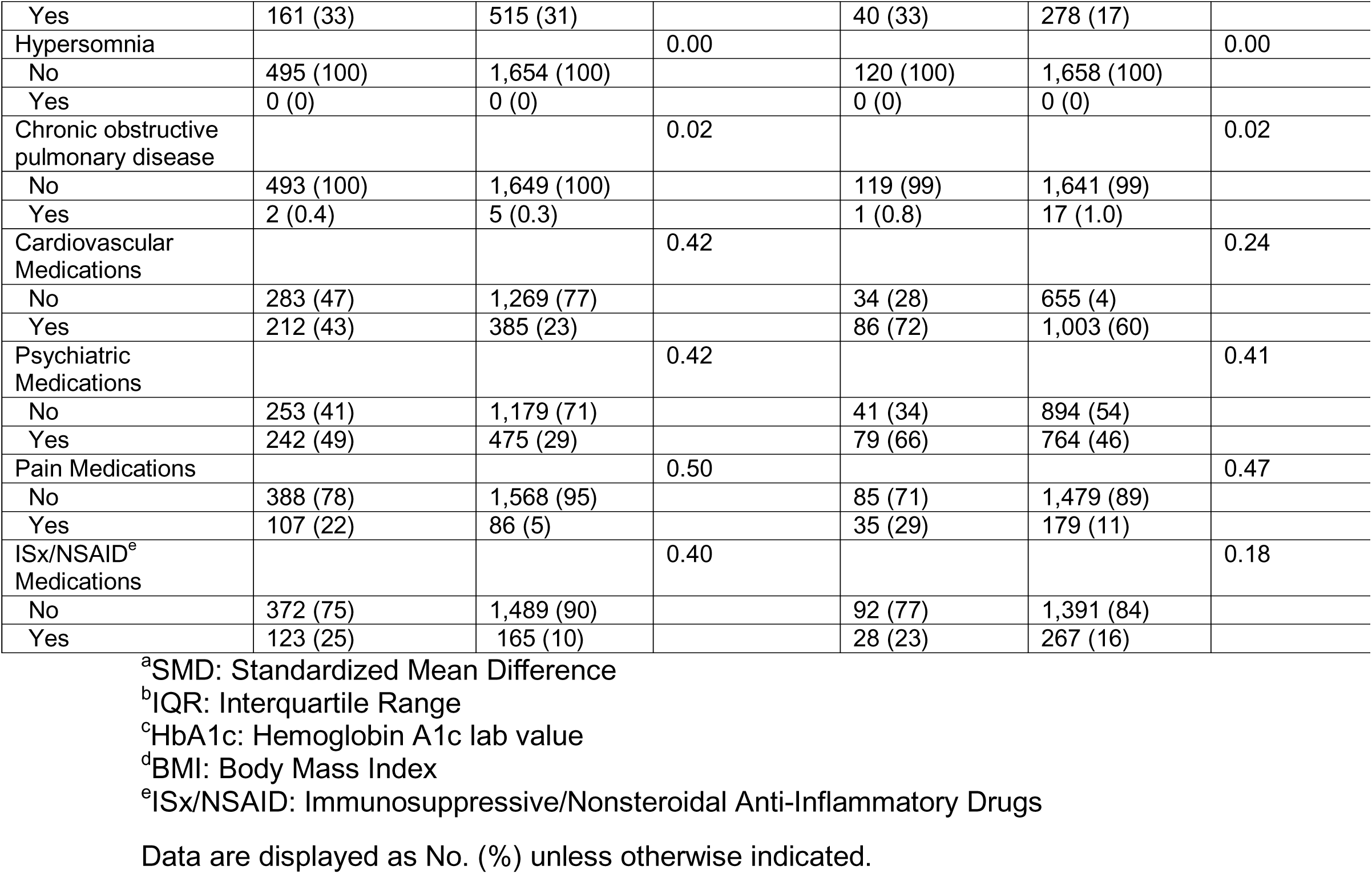
Patient characteristics.

| Characteristic | Nondiabetic patients |  |  | Diabetic patients |  |  |
| --- | --- | --- | --- | --- | --- | --- |
|  | Semaglutide (N=495) | NC (N=1,654) | SMD<sup>a</sup> | Semaglutide (N=120) | NC (N=1,658) | SMD<sup>a</sup> |
| Age, median (IQR)<sup>b</sup> | 45 (37, 53) | 46 (35, 57) | 0.09 | 51 (43, 57) | 57 (46, 66) | 0.47 |
| Sex |  |  | 0.06 |  |  | 0.41 |
| Female | 374 (76) | 1,206 (73) |  | 91 (76) | 942 (57) |  |
| Male | 121 (24) | 448 (27) |  | 29 (24) | 716 (43) |  |
| Race |  |  | 0.24 |  |  | 0.41 |
| Black or African American | 66 (13) | 110 (6.7) |  | 27 (23) | 156 (9.4) |  |
| White | 382 (77) | 1,421 (86) |  | 82 (68) | 1,357 (82) |  |
| Unknown | 18 (3.6) | 47 (2.8) |  | 5 (4.2) | 27 (1.6) |  |
| Other | 29 (5.9) | 76 (4.6) |  | 6 (5.0) | 118 (7.1) |  |
| HbA1c<sup>c</sup> |  |  | 0.09 |  |  | 0.93 |
| < 5.7 | 232 (82) | 843 (79) |  | 44 (44) | 165 (10) |  |
| 5.7 - 6.4 | 50 (18) | 221 (21) |  | 25 (25) | 349 (22) |  |
| 6.5 and above | 0 (0) | 1 (<0.1) |  | 30 (30) | 1,083 (68) |  |
| Unknown | 213 | 589 |  | 21 | 61 |  |
| Initial BMI<sup>d</sup> | 39 (34, 43) | 40 (35, 46) |  | 40 (34, 46) | 40 (35, 46) |  |
| Acute Care Visits, median (IQR)<sup>b</sup> | 1.0 (0.0, 3.0) | 1.0 (0.0, 3.0) | 0.15 | 2 (1, 5) | 3 (1, 8) | 0.26 |
| Ambulatory Care Visits, median (IQR)<sup>b</sup> | 91 (40, 185) | 143 (64, 295) | 0.45 | 214 (84, 348) | 353 (172, 810) | 0.65 |
| Obstructive Sleep Apnea |  |  | 0.20 |  |  | 0.09 |
| No | 403 (81) | 1,208 (73) |  | 73 (61) | 1,078 (65) |  |
| Yes | 92 (19) | 446 (27) |  | 47 (39) | 580 (35) |  |
| Myocardial Infarction |  |  | 0.00 |  |  | 0.19 |
| No | 492 (99) | 1,644 (99) |  | 119 (99) | 1,597 (96) |  |
| Yes | 3 (0.6) | 10 (0.6) |  | 1 (0.8) | 61 (3.7) |  |
| Stroke |  |  | 0.07 |  |  | 0.12 |
| No | 495 (100) | 1,650 (100) |  | 120 (100) | 1,647 (99) |  |
| Yes | 0 (0) | 4 (0.2) |  | 0 (0) | 11 (0.7) |  |
| Heart Failure |  |  | 0.12 |  |  | 0.32 |
| No | 495 (100) | 1,642 (99) |  | 120 (100) | 1,579 (95) |  |
| Yes | 0 (0) | 12 (0.7) |  | 0 (0) | 79 (4.8) |  |
| Peripheral Procedures |  |  | 0.04 |  |  | 0.10 |
| No | 494 (100) | 1,653 (100) |  | 119 (99) | 1,656 (100) |  |
| Yes | 1 (0.2) | 1 (<0.1) |  | 1 (0.8) | 2 (0.1) |  |
| Hypertension |  |  | 0.03 |  |  | 0.04 |
| No | 306 (62) | 1,045 (63) |  | 33 (28) | 429 (26) |  |
| Yes | 189 (38) | 609 (37) |  | 87 (73) | 1,229 (74) |  |
| Atrial fibrillation |  |  | 0.04 |  |  | 0.03 |
| No | 486 (98) | 1,614 (98) |  | 115 (96) | 1,600 (97) |  |
| Yes | 9 (1.8) | 40 (2.4) |  | 5 (4.2) | 58 (3.5) |  |
| Anxiety disorder |  |  | 0.13 |  |  | 0.01 |
| No | 491 (99) | 1,613 (98) |  | 112 (93) | 1,545 (93) |  |
| Yes | 4 (0.8) | 41 (2.5) |  | 8 (6.7) | 113 (6.8) |  |
| Chronic kidney disease |  |  | 0.03 |  |  | 0.39 |
| No | 334 (67) | 1,139 (69) |  | 80 (67) | 1,380 (83) |  |
| Yes | 161 (33) | 515 (31) |  | 40 (33) | 278 (17) |  |
| Hypersomnia |  |  | 0.00 |  |  | 0.00 |
| No | 495 (100) | 1,654 (100) |  | 120 (100) | 1,658 (100) |  |
| Yes | 0 (0) | 0 (0) |  | 0 (0) | 0 (0) |  |
| Chronic obstructive pulmonary disease |  |  | 0.02 |  |  | 0.02 |
| No | 493 (100) | 1,649 (100) |  | 119 (99) | 1,641 (99) |  |
| Yes | 2 (0.4) | 5 (0.3) |  | 1 (0.8) | 17 (1.0) |  |
| Cardiovascular Medications |  |  | 0.42 |  |  | 0.24 |
| No | 283 (47) | 1,269 (77) |  | 34 (28) | 655 (4) |  |
| Yes | 212 (43) | 385 (23) |  | 86 (72) | 1,003 (60) |  |
| Psychiatric Medications |  |  | 0.42 |  |  | 0.41 |
| No | 253 (41) | 1,179 (71) |  | 41 (34) | 894 (54) |  |
| Yes | 242 (49) | 475 (29) |  | 79 (66) | 764 (46) |  |
| Pain Medications |  |  | 0.50 |  |  | 0.47 |
| No | 388 (78) | 1,568 (95) |  | 85 (71) | 1,479 (89) |  |
| Yes | 107 (22) | 86 (5) |  | 35 (29) | 179 (11) |  |
| ISx/NSAID<sup>e</sup> Medications |  |  | 0.40 |  |  | 0.18 |
| No | 372 (75) | 1,489 (90) |  | 92 (77) | 1,391 (84) |  |
| Yes | 123 (25) | 165 (10) |  | 28 (23) | 267 (16) |  |

<sup>a</sup>SMD: Standardized Mean Difference
<sup>b</sup>IQR: Interquartile Range
<sup>c</sup>HbA1c: Hemoglobin A1c lab value
<sup>d</sup>BMI: Body Mass Index
<sup>e</sup>ISx/NSAID: Immunosuppressive/Nonsteroidal Anti-Inflammatory Drugs
Data are displayed as No. (%) unless otherwise indicated.

### Study Outcomes

The study aimed to evaluate achievement and time to achievement of 10% reduction in body weight as the primary outcome measure. Percent weight loss was calculated as 100% *(weight – baseline weight) / baseline weight. After their baseline weight measurement, weight measurements after initiating semaglutide or lifestyle counseling were obtained from the EHR. The time in days for participants to reach a weight reduction of 10% from their baseline weight was obtained and utilized in a time-to-event analysis.^14^ Individuals who failed to achieve a 10% weight loss were considered right-censored. The censor date was calculated as the earlier of either the termination of therapy, most recent weight measurement present in the EHR, or 2 years following therapy initiation. If instances of bariatric surgery, cancer, or pregnancy occurred after semaglutide or lifestyle counseling initiation, the first date recorded for these events was also categorized as censoring events. This approach ensured a comparable tracking duration for participants in the semaglutide and lifestyle counseling groups, given the recent approval of semaglutide for weight loss in 2021, which limited the to track individuals for an extended period. A complete diagram illustrating the study design timeline is available in eFigure1.

### Statistical Analysis

Data were collected from 2008 to 2023 for individuals undergoing lifestyle counseling and from 2018 to 2023 for those receiving semaglutide treatment. Baseline information encompassed clinical, demographic, and health service utilization variables. Categorical data were presented with absolute and relative frequencies, while continuous data were summarized with mean and standard deviation (SD) metrics. Standardized mean differences (SMD) across exposure groups were reported for all covariates. Propensity score matching^15^ was employed to balance baseline covariates between the two treatment groups, with 1:1 matching conducted in order estimate the average treatment effect on the treated (ATT) for the semaglutide cohort compared to lifestyle counseling. Kaplan Meier curves^16,17^ and Cox proportional hazards models^18^ were fit on the matched dataset. Hazard ratios are reported to estimate the hazard of achieving 10% weight loss between semaglutide and behavior counseling. E-values^19^ were calculated to measure the amount of unmeasured confounding that would be needed to nullify the association, providing a measure of robustness for the observational analysis.

## Results

### Study Population

From 21,503 individuals with any exposure to semaglutide, 3,352 had prescriptions longer than 16 weeks needed to escalate the semaglutide dosage and were included for study. Of these, 120 were classified using phenotype definitions as having type 2 diabetes mellitus and 495 were classified as non-diabetic. For lifestyle counseling alone, 262,699 had at least 1 session scheduled. Of these, 3,312 were eligible for study with 1,658 having type 2 diabetes mellitus and 1,654 without type 2 diabetes mellitus.

Baseline characteristics of clinical variables, demographic variables, and health service utilization variables were gathered for both diabetic and non-diabetic populations and are presented in Table 1. Baseline characteristics show generally good balance among populations. The majority of semaglutide patients were female, and the majority of patients in groups were white and had low rates of clinical diagnoses screened for in the table.

### Propensity Score Analysis

Propensity score matching was employed to achieve covariate balance when comparing treatments for both the non-diabetic and diabetic populations. This involved a 1:1 nearest neighbor matching using logistic regression modeling of propensity scores,^20^ which was based on the odds of receiving semaglutide or lifestyle behavior therapy. These scores were derived from baseline covariates including clinical, demographic, and health service utilization variables. This matching technique was selected due to a larger pool of patients undergoing lifestyle counseling, enabling the estimation of the Average Treatment Effect on the Treated (ATT).^21^ Propensity score matching calculations were done using the R programming language^22^ and the MatchIt package.^23^ Covariate balance plots (i.e., ‘love plots’) before and after matching are available in eFigure2 and eFigure3.

### Achievement of 10% Weight Loss

In the nondiabetic population, 506/1,654 (30.6%) of individuals in the lifestyle counseling alone group lost 10% of their initial weight, compared with 222/495 (44.8%) of individuals in the semaglutide group over the 2 year study time frame (Relative Risk 1.47; 95% CI, 1.30-1.66). In the diabetic population, 369/1,658 (22.2%) of individuals in the lifestyle counseling alone group lost 10% of their initial weight compared with 45/120 (37.5%) of individuals in the semaglutide group (Relative Risk 1.69; 95% CI, 1.32-2.16). For the 5% weight loss benchmark, 327/495 (66.1%) patients in the non-diabetic semaglutide group compared with 942/1,654 (56.9%) patients in the lifestyle counseling alone group lost 5% of weight (Relative Risk 1.15; 95% CI, 1.07-1.24). In diabetic populations, 64/120 (53.3%) who received semaglutide compared with 830/1,658 (50.0%) who received lifestyle counseling alone lost 5% weight (Relative Risk 1.06; 95% CI, 0.90-1.27).

Following covariate matching using propensity scores, Kaplan-Meier curves were generated to estimate survival probability for achieving a 10% weight reduction (Figure 3). The Kaplan-Meier curves were constructed with time to achievement of 10% weight loss as the outcome, measured in days, and applied to the matched cohorts comprising of 120 patients each in the semaglutide and lifestyle counseling alone groups for diabetic patients, and 495 patients for non-diabetic patients.

**Figure 1:**
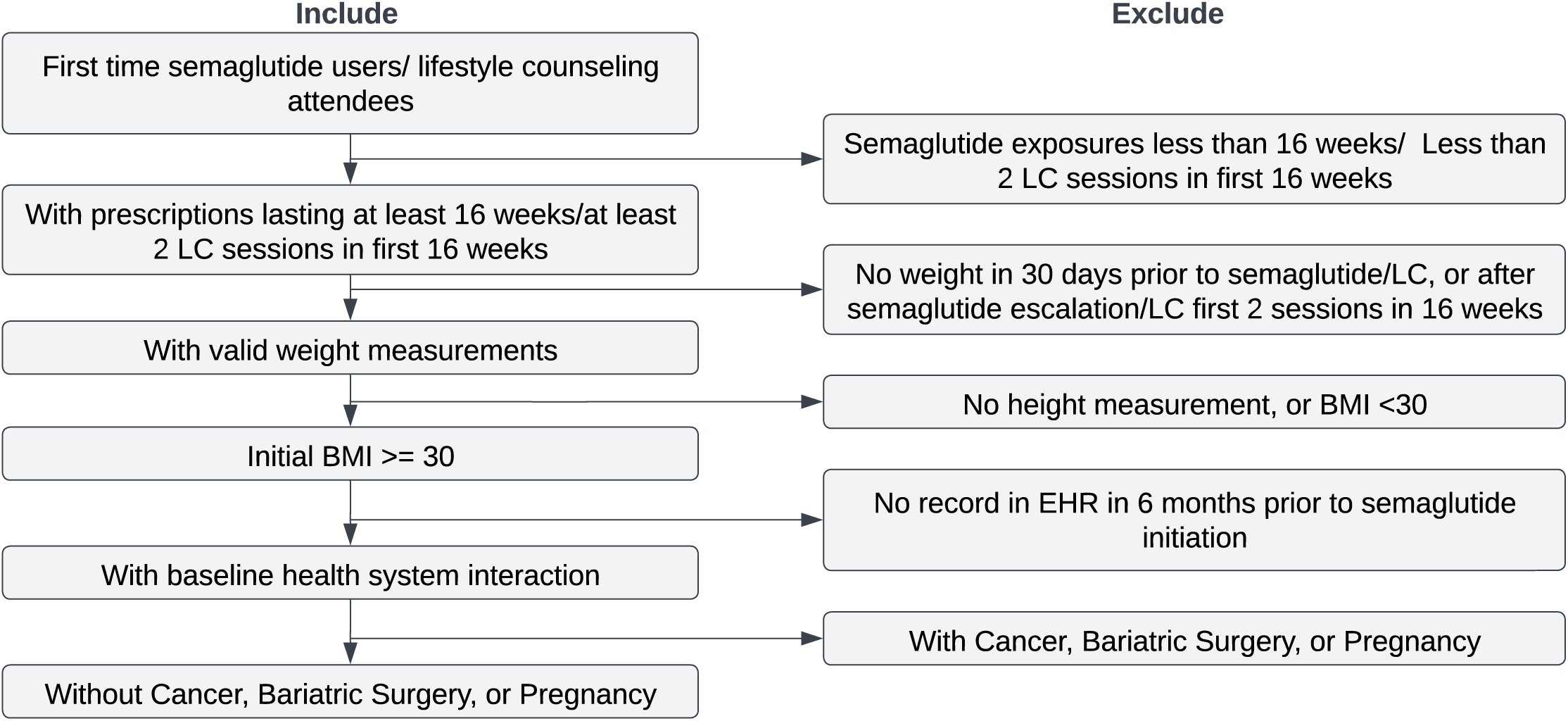
Patient Selection Criteria. Beginning with any individual with a present electronic health record of semaglutide prescription or lifestyle counseling, this diagram described the inclusion and exclusion criteria to arrive at the analytic cohorts. Abbreviations: LC: Lifestyle Counseling, BMI, Body Mass Index, EHR, Electronic Health Record

**Figure 2:**
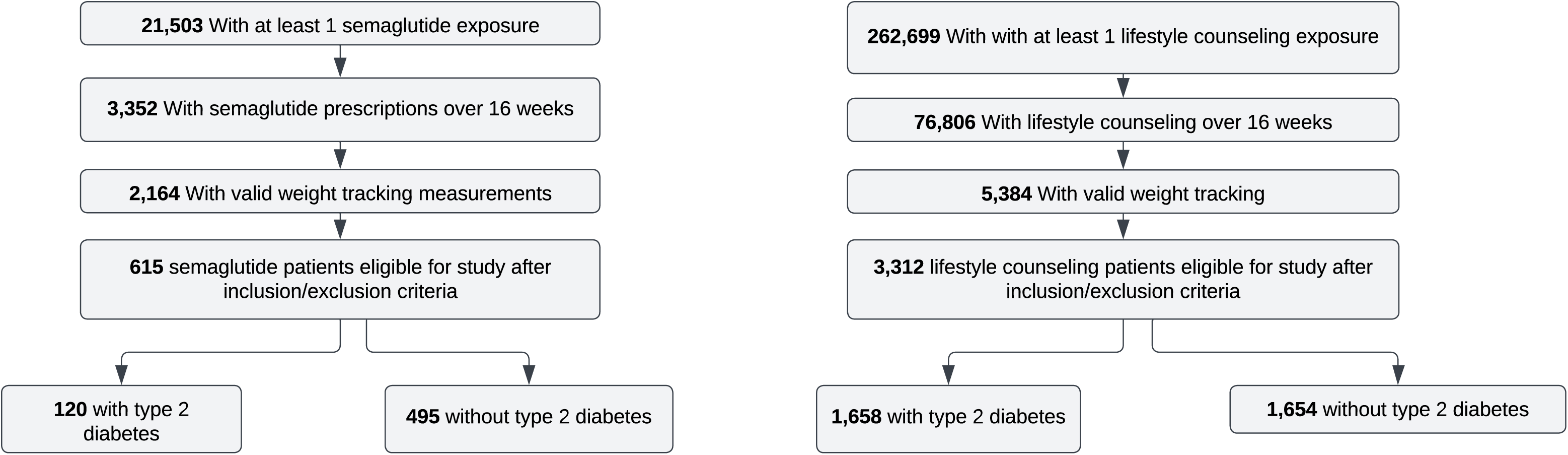
Patient Flowchart. Valid weight tracking requires a baseline weight in the 30 days prior to therapy initiation and one following 16 weeks of exposure to semaglutide or lifestyle counseling.

**Figure 3:**
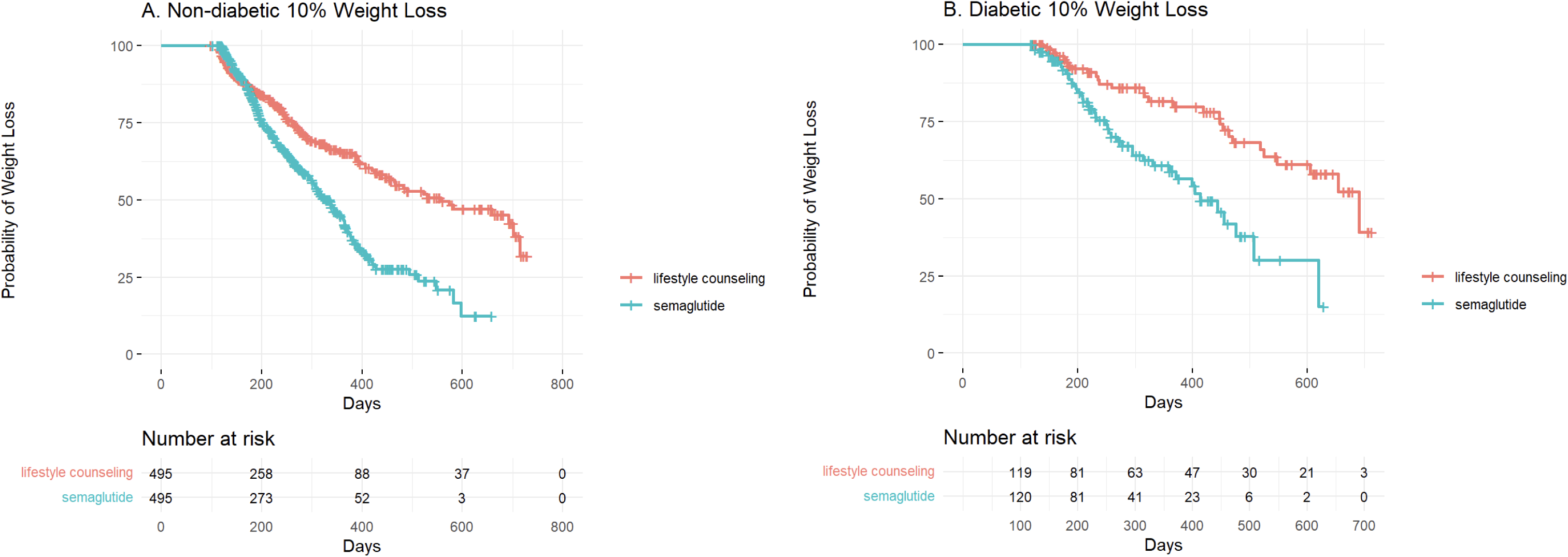
10% Weight Loss Kaplan-Meier Curves. A, Kaplan Meier curves for 10% weight loss in the non-diabetic population, B, Kaplan Meier curves for 10% weight loss in the diabetic population.

In the non-diabetic subgroup, both participants receiving semaglutide and lifestyle counseling alone demonstrated notable efficacy in achieving the 10% weight loss target. However, significant differences were observed when comparing survival curves between intervention (log rank <0.001, Figure 3). Hazard ratios were obtained from a fitted Cox-Regression model^18^ and revealed that individuals on semaglutide had significantly greater hazards of achieving 10% weight loss (1.63 [95% CI: 1.32, 2.01; p<0.001]) when compared to nutrition counseling.

Similar results were observed in the population of patients with diabetes, with large differences between semaglutide and lifestyle counseling weight loss survival curves (log rank <0.001, Figure 3). Hazard ratios were also calculated from a fitted Cox-Regression model and revealed that diabetic individuals on semaglutide also had significantly greater hazards of achieving 10% weight loss (2.45 [95% CI: 1.47, 4.09; p<0.001]) when compared to nutrition counseling, with larger effects then among non-diabetic individuals.

Sensitivity analysis was conducted using E values^24^ in order to determine the amount of unmeasured confounding that would need to occur on order to change the interpretation of the results. Hazard ratios and 95% confidence intervals were calculated into risk ratios, which were then used to calculate the E-value. For the non-diabetic population, the E-value was 2.38 [95% CI: 1.91, 2.91], and for the diabetic population it was 3.35 [95% CI: 2.21, 4.82]. These measures suggest that unmeasured confounding with very large effects would have to exist in order to nullify associations.

## Discussion

By leveraging a robust causal inference approach, the current study suggests that individuals taking semaglutide had greater success at achieving 10% weight loss when compared to those that received lifestyle counseling alone. Results were observed among patients with or without type 2 diabetes, with greater effects among those with this condition. Often, in diabetic individuals the medical lifestyle therapy is intended to prevent more weight gain rather than achieve weight loss in addition to glycemic control^5^, individuals taking semaglutide see improved weight loss outcomes in addition to glycemic control not seen with lifestyle therapy, holding a host of potential positive outcomes including decreased risk of cardiovascular events.^25^

This study has several strengths. While previous randomized clinical trials have showed the effectiveness of semaglutide under similar conditions,^26^ other observational studies have focused only on 2.0mg and not 2.4mg dosages of semaglutide,^27^ individuals with type 2 diabetes,^27,28^ or used synthetic data.^29^ Our study provides novel insight into both diabetic and non-diabetic treatment groups taking 2.4mg semaglutide in a large, multi-site population across the central United States with real-world data. In addition, the rich EHR data source was used to extract clinical factors that were used in a causal inference approach that provided robustness. There are also important limitations. The study lacks information regarding socioeconomic factors and actual lifestyle variables that may affect the ability for individuals receiving therapies to lose weight. Potentially related to the significant reduction in number of patients with at least 16 weeks exposure relative to initial prescribing (3,352 and 21,503 respectively), semaglutide incurs a high cost for patients,^30^ and likely a significantly higher cost for patients than lifestyle counseling^31^ or bariatric surgery.^29^ Socioeconomic status could have an effect on the analysis, as low and middle income groups often carry the greatest weight of the obesity burden. Future comparative effectiveness studies that link individual level socioeconomic characteristics as well as out of pocket patient costs are warranted to incorporate patient centered economic outcomes.^32^

Strong adherence and lifestyle information is also lacking from the dataset and could be a limitation. Though individuals in the lifestyle counseling group receiving information during sessions, their adherence to plans and strategies for changing diet and exercise may vary. Likewise, semaglutide exposure for this study was based upon prescribing activity. Outpatient pharmacy dispensing may provide a greater sense of adherence in subsequent studies. Moreover, the actual diet and exercise patterns of individuals receiving semaglutide and lifestyle counseling would provide further information to help minimize unbalance between exposure groups. Additionally, incorporating both lifestyle counseling and semaglutide together for weight loss therapy can be studied for additive effects. Future studies can incorporate longitudinal data to better evaluate long-term efficacy and sustainability of semaglutide versus lifestyle counseling. While this study was limited in its window of observations to 2 years, studies with long-term data could look to study effects such as prevalence of weight regain or successful long-term weight loss. Longitudinal data would also allow study for adherence rates, as participants in the semaglutide group had high rates of patients that did not complete more than 16 weeks of prescriptions. This is due to an indeterminate effect that may be due partially to tolerability issues with mechanisms of semaglutide or with a lack of new data on a recently approved medication. Finally, variation in treatment mechanisms and effect between semaglutide and lifestyle and their impact on different underlying populations warrants further considerations. Pharmacological interventions biologically effect patients in different ways than behavior based interventions, such as how semaglutide changes appetite regulation, gastric emptying and blood glucose stabilization.^33^ These effects may be more or less desirable depending on the treatment population, which depends on factors such as diabetes incidence. Personalized approaches could look to incorporate these factors as well as genetic, lifestyle, and metabolic factors to determine best therapy.

## Conclusions

After adjusting for confounding, our data demonstrate that individuals taking semaglutide had greater success at achieving 10% weight loss than individuals receiving lifestyle counseling alone. While there are many factors that impact the decision to initiate adjunct semaglutide therapy versus traditional lifestyle counseling alone, these real-world data supports the conclusion that semaglutide is more effective for weight loss than lifestyle counseling alone in an obese population.

## Data Availability

The data used in this study were derived from electronic health records (EHRs) and contain sensitive patient information. Due to legal, ethical, and institutional restrictions related to patient privacy and confidentiality (including HIPAA regulations), these data cannot be shared publicly. Access to the raw EHR data is therefore not available.

## Author Contributions

*Concept and Design:* Will Powell, Diego Mazzotti, Stephen Herrman, Lemuel R. Waitman

*Acquisition and Analysis of Data:* Will Powell

*Drafting of Manuscript:* Will Powell *Critical Review of Manuscript:* All Authors *Statistical Analysis:* Will Powell

*Obtaining Funding:* Lemuel R. Waitman

*Administrative Support/Supervision:* Lemuel R. Waitman

