## Supplementary Files for "Comparison of semaglutide and lifestyle counseling for weight loss using electronic health records"

**Supplementary Information Contents:**

**eFigure 1.** Overview of the study design timeline

**eFigure 2.** Non-diabetic population Propensity Score Matching Love Plot

**eFigure 3.** Diabetic population Propensity Score Matching Love Plot

**eFigure 1.** Overview of the study design timeline

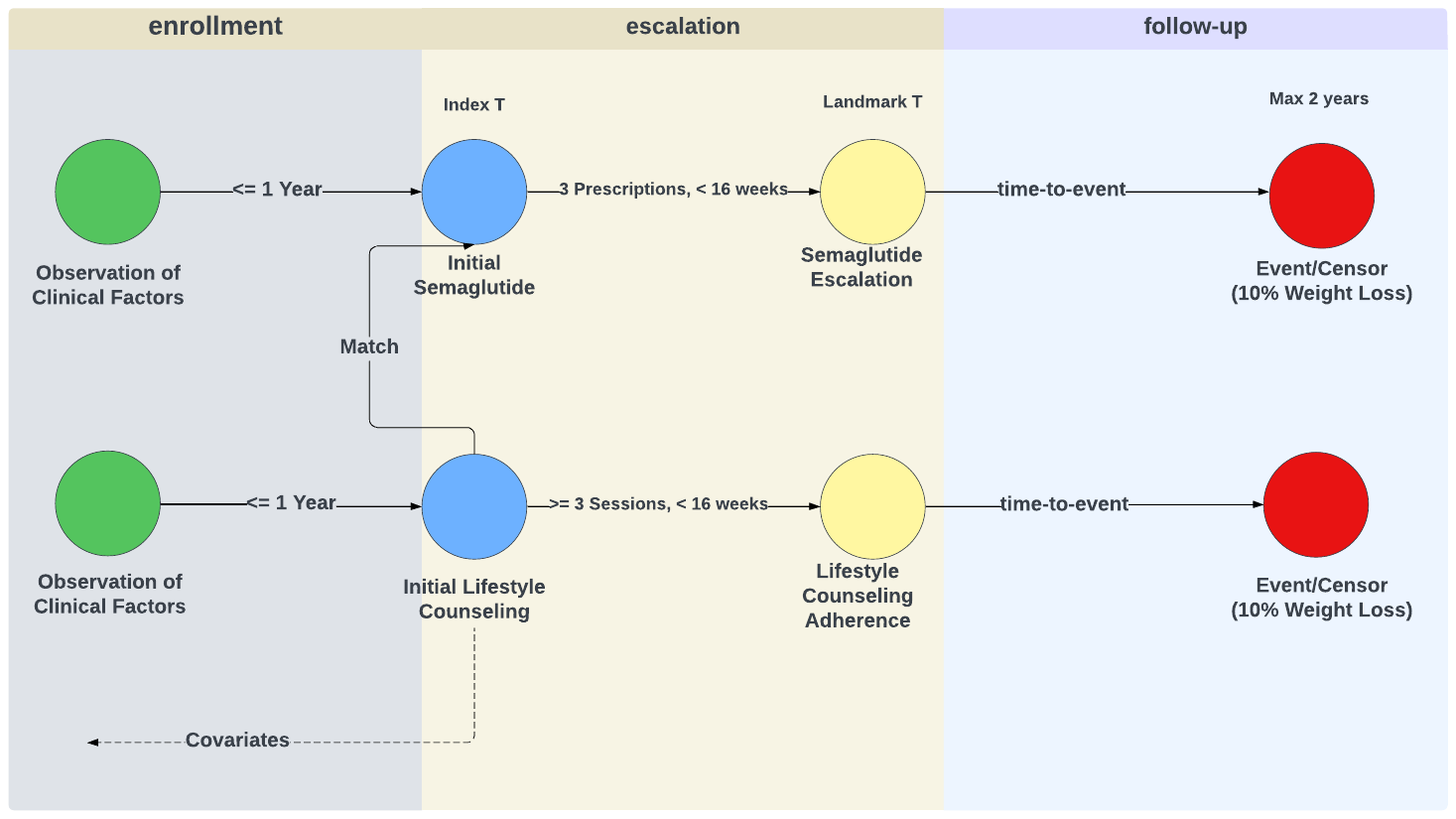

eFigure1 shows the study design timeline. At Index T, participants begin either semaglutide or nutrition counseling, while at Landmark T they reach semaglutide prescriptions lasting 16 weeks or have at least 3 nutrition counseling sessions in the first 16 weeks of their treatment.

**eFigure 2.** Non-diabetic population Propensity Score Matching Love Plot

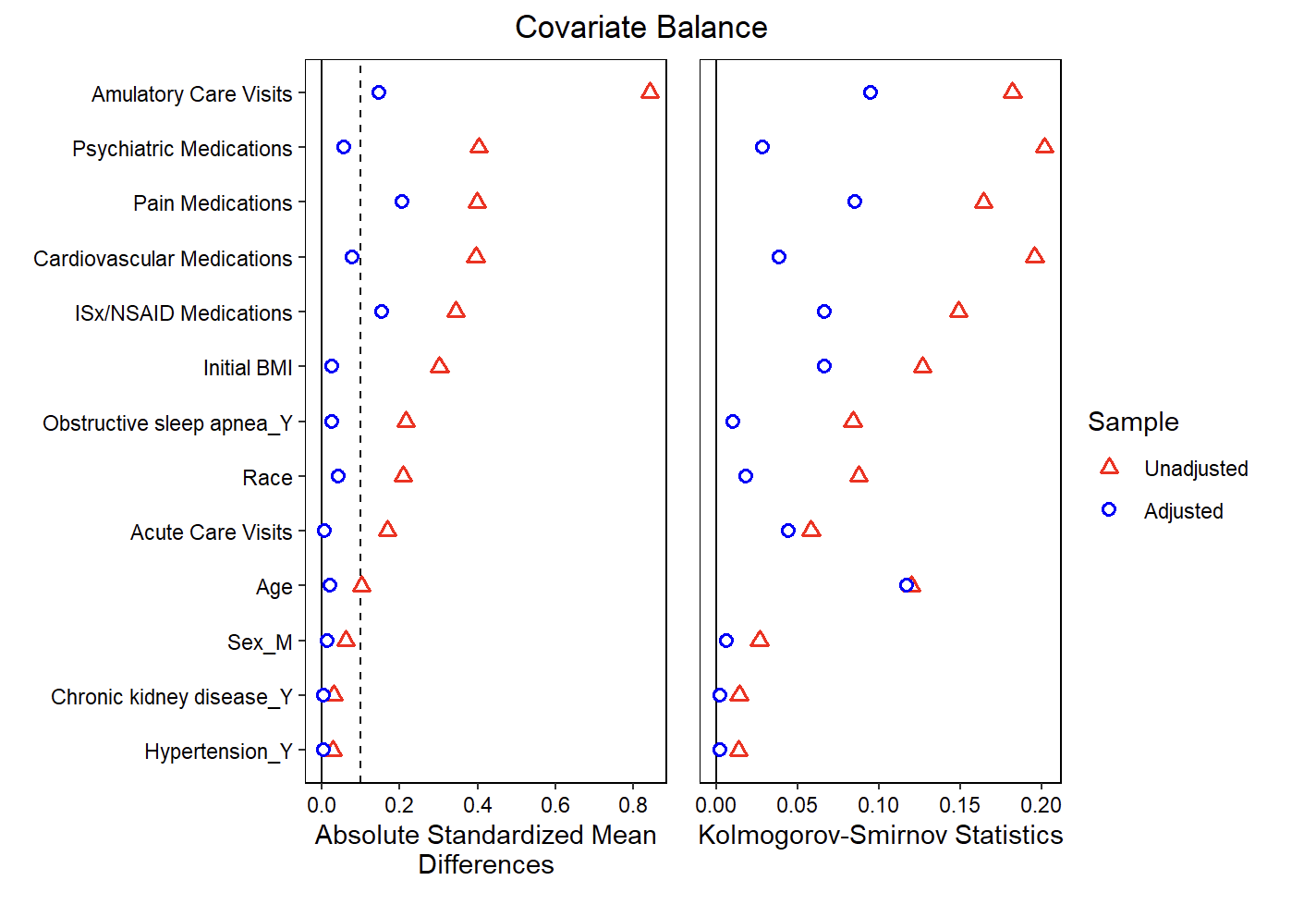

eFigure2 shows covariate balances before and after propensity score matching in the non-diabetic cohorts. Relevant covariate groups show good balance with low Standardized Mean Differencess after propensity score matching. Abbreviations: ISx/NSAID, Immunosuppressive/Nonsteroidal Anti-Inflammatory Drugs

**eFigure 3.** Diabetic population Propensity Score Matching Love Plot

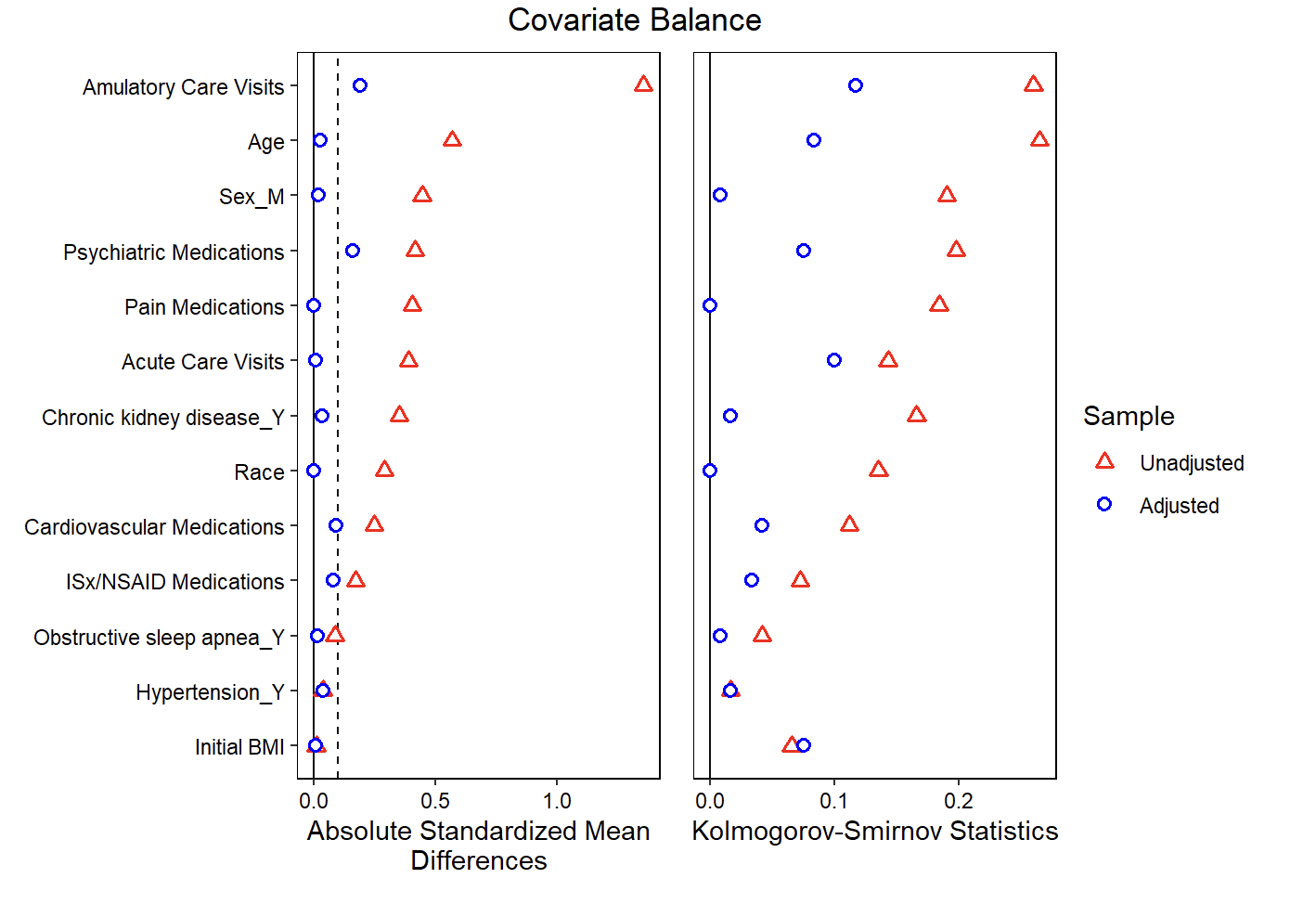

eFigure3 shows the covariate balances before and after propensity score matching in the diabetic cohorts. Relevant covariate groups show good balance with low SMDs after propensity score matching. Abbreviations: ISx/NSAID, Immunosuppressive/Nonsteroidal Anti-Inflammatory Drugs

### Primary Outcomes

| Outcome | Description | Specifications |
| --- | --- | --- |
| 10% Weight Loss | Recorded as first incidence of achieving 10% weight loss on semaglutide  Calculated as (weight – initial weight) / initial weight * 100% | A event/censor variable |

### Primary Exposures

| Exposure | Description | Specifications |
| --- | --- | --- |
| Semaglutide | Type of semaglutide dose received  Wegovy RxNorm codes: 2553503, 25536022553608, 2553902, 2554103, 2553506, 2553603, 2553803, 2553903, 2554104, 2553501, 2553601, 2553802, 2553901, 2554102, 2553400, 2553600, 2553606, 2553900,2554101  Ozempic RxNorm codes: 2619153, 1991308,2599364, 2619154,2398842, 1991311, 1991317,2599365, 1991306,2619152, 1991316, 2398841, 2599362, 1991303, 2619151, 2599361, 2398844, 1991309, 1991305, 2398843  Rybelsus RxNorm codes: 2200653, 2200657, 2200646, 2200654, 2200658, 2200650, 2200652, 2200656, 2200644, 2200651,2200655, 2200640, 2200643, 2200642, 2200641, 2200647 | Categorical Variable with  Wegovy (semaglutide injection 2.4 mg)  Ozempic (semaglutide injection 0.5mg, 1.0mg, or 2.0 mg)  Rybelsus (semaglutide tablets 7mg or 14mg) |
| Nutrition Behavior Therapy | HCPCS codes: G0270, G0271, G0447, 97802, 97803, 97804, G0473 | Binary variable |

### Demographic Variables

| Variable | Description | Specifications |
| --- | --- | --- |
| patid | Patient Identification number | A nominal variable |
| Age | Patient age at start of semaglutide exposure | A continuous variable |
| Sex | Patient coded sex variable | Categorical Variable with  1= male  0=female |
| Race | Patients identified racial identity | Categorical Variable with  01=American Indian or Alaska Native  02=Asian  03=Black or African American  04=Native Hawaiian or Other Pacific Islander  05=White 06=Multiple race  07=Refuse to answer  NI=No information  UN=Unknown  OT=Other |
| Type 2 diabetes mellitus | Diagnosis of Type 2 diabetes mellitus is used to stratify into two distinct groups for study. See appendix for description of codes used. | Categorical Variable with  1= diagnosis of type 2 diabetes mellitus  0=no diagnosis of type 2 diabetes mellitus |

### Clinical Variables

| Variable | Description | Specifications |
| --- | --- | --- |
| Obstructive sleep apnea | Incidence in year prior through escalation date | ICD-9-CM: 327.20, 327.23, 327.29, 780.51, 780.53, 780.57  ICD-10-CM: G47.30, G47.33, G47.39 |
| Myocardial infarction | Incidence in year prior through escalation date | ICD-9-CM: 410.X, 412.X  ICD-10-CM: I21.X, I22.X, I23.X |
| Stroke | Incidence in year prior through escalation date | ICD-9-CM: 431.X, 434.X  ICD-10-CM: I61.X, I62.X, I64.X |
| Heart failure | Incidence in year prior through escalation date | ICD-9-CM: 428.X  ICD-10-CM: I50.X |
| Cardiac revascularization | Incidence in year prior through escalation date | ICD-9-CM: 327.20, 327.23, 327.29, 780.51, 780.53, 780.57  ICD-10-PCS: 021X, 027X  HCPCS: 92920-92944, 92973, 92974, 92975, 92980, 92981, 92982, 92984, 92995, 92996 |
| Peripheral procedures | Incidence in year prior through escalation date | ICD-9-CM: 00.4, 00.66  HCPCS: 37220-37235, 37215-37218, 37236-37249, 37211-37214, 37184-37188 |
| Hypertension | Incidence in year prior through escalation date | ICD-9-CM: 401.X, 402.X, 403.X, 404.X, 405.X  ICD-10-CM: I10.X, I11.X, I12.X, I13.X, R03.X |
| Atrial fibrillation | Incidence in year prior through escalation date | ICD-9-CM: 427.3  ICD-10-CM: I48.X |
| Anxiety disorder | Incidence in year prior through escalation date | ICD-9-CM: 300.X  ICD-10-CM: N18.X |
| Chronic kidney disease | Incidence in year prior through escalation date | ICD-9-CM: 585.X  ICD-10-CM: F40.X-F48.X |
| Hypersomnia | Incidence in year prior through escalation date | ICD-9-CM: 327.1  ICD-10-CM: F51.1, G47.1 |
| Insomnia | Incidence in year prior through escalation date | ICD-9-CM: 327.0, 307.41, 307.42, 780.52  ICD-10-CM: F51.01, F51.02, F51.09 |
| Chronic obstructive pulmonary disease | Incidence in year prior through escalation date | ICD-9-CM: 496.X  ICD-10-CM: J44.X |
| HbA1c | Initial Hemoglobin A1c lab value occurring in the 30 days prior to semaglutide initiation | LOINC: 4548-4, 41995-2, 55454-3, 71875-9, 549-2, 17856-6, 59261-8, 62388-4, 17855-8 |
| Medications | List of relevant medications | See link |

Abbreviations: ICD: International Classification of Diseases; CM: Clinical Modification; CPT: Current Procedural Terminology; PCS: Procedure Coding System; HCPCS: Healthcare Common Procedure Coding System

Medications List See: <https://raw.githubusercontent.com/RWD2E/phecdm/main/res/valueset_autogen/ecqm-medication.json>

### Health Service Utilization Variables

| Variable | Description | Specifications |
| --- | --- | --- |
| sema_counts | Number of semaglutide prescriptions recorded in EHR | Taken from Wegovy, Ozempic, or Rybelsus RxNorm codes and PCORI CDM PRESCRIBING table |
| num_weights | Number of weights measurements recorded in EHR | Obtained from PCORI CDM VITAL table |
| encounter_ac | Number of total visits in health system that are of type Acute Care | PROVIDER  ENC_TYPE:  ED=Emergency Department  EI=Emergency Department Admit to Inpatient Hospital Stay (permissible substitution) IP=Inpatient Hospital Stay |
| encounter_av | Number of total ambulatory visits in EHR | PROVIDER  ENC_TYPE:  AV=Ambulatory Visit  IS=Non-Acute Institutional Stay  IC=Institutional Professional Consult (permissible substitution)  TH=Telehealth  OA=Other Ambulatory Visit |
| Insurance_type | Type of insurance patient enrolled with |  |

### Censoring/Exclusion Variables

| Variable | Description | Specifications |
| --- | --- | --- |
| Bariatric surgery | Recorded bariatric surgery procedure occurring at any time prior to semaglutide initiation through escalation (exclusion) or after semaglutide escalation (censor) | ICD-9: 43.82, 43.89, 44.3, 44.31, 44.38, 44.39, 44.68, 44.95, 44.96, 44.97, 44.99, 44.5, 45.51, 45.9  ICD-10: Z98.0 |
| Pregnancy | Recorded pregnancy occurring in the year prior to semaglutide initiation through escalation (exclusion) or after semaglutide escalation (censor) | ICD-9: 630-679  ICD-10: Ox |
| Cancer | Recorded malignant cancer occurring in the year prior to semaglutide initiation through escalation (exclusion) or after semaglutide escalation (censor) | ICD-9: 140-215, 217-240  ICD-10: C, D  Exclusions:  ICD-10: D22 |

### Type 2 diabetes mellitus phenotype

The method of phenotyping for incidence of type 2 diabetes mellitus was derived from the method used by Wiese et. Al [cite] where a patient is diagnosed with incidence of type 2 diabetes mellitus if they have (1) a coded inpatient or outpatient T2DM diagnosis (ICD9/ICD10) (Table 1) and an antidiabetic medication prescription (Table 2) within the 90 days following the diagnosis date (CP1), (2) a coded T2DM diagnosis and an outpatient glycolated hemoglobin (HbA1C) value ≥6.5% within 90 days before or after the diagnosis date (CP2) (laboratory values queried using Logical Observation Identifiers, Names and Codes (LOINC) codes (Table 3), or (3) any antidiabetic medication prescription within 90 days before or after an outpatient HbA1C value ≥6.5% (CP3)

**Table 1.** List of ICD9 and ICD10 codes for the identification of patients with Type 2 diabetes using coded diagnoses

| Classification | Code list | Codes |
| --- | --- | --- |
| Qualifying coded diagnosis for Type 2 diabetes | ICD9 | 250.00, 250.2, 250.22, 250.3, 250.32, 250.4, 250.42, 250, 250.02, 250.5, 250.52, 250.6, 250.62, 250.7, 250.72, 250.8, 250.82, 250.9, 250.92 |
|  | ICD10 | E11.00, E11.01, E11.21, E11.22, E11.29, E11.311, E11.319, E11.321, E11.329, E11.331, E11.339, E11.341, E11.349, E11.351, E11.359, E11.36, E11.39, E11.40, E11.41, E11.42, E11.43, E11.44, E11.49, E11.51, E11.52, E11.59, E11.610, E11.618, E11.620, E11.621, E11.622, E11.628, E11.630, E11.638, E11.641, E11.649, E11.65, E11.69, E11.8, E11.9 |
| Exclusion criteria |  |  |
| Impaired fasting glucose; Impaired glucose tolerance; Other unspecified Diabetes | ICD9 | 790.21, 790.22, 790.29, 791.5, 277.7, 790.29 |
|  | ICD10 | R73.01, R73,02, R73.0, R81.*, E88.81, Z13.1, E13.*, E08.*, E09.* |
| Type 1 diabetes | ICD9 | 250.03 |
|  | ICD10 | E10.*, |
| Pregnancy/gestational diabetes | ICD9 | 648.8 |
|  | ICD10 | O24.*, |

*Indicates all codes end with numeric value 0-9 are included

**Table 2.** List of proprietary and generic medication names used to identify patients with Type 2 diabetes^1^

| Medication class | Generic names [proprietary names] |
| --- | --- |
| Sulfonylurea | tolazamide [Tolinase]; acetohexamide [Dymelor]; tolbutamide [Orinase, Orinase Diagnostic, Tol-Tab]  chlorpropamide [Diabinese] [diabanase][diabinase]; glipizide [Glucotrol, Glucotrol XL, Glucatrol];  glyburide [Micronase, Glynase, Diabetamide, Diabeta]; glimepiride [Amaryl] |
| Biguanide | metformin [Glucophage] |
| Meglitanide | repaglinide [Prandin]; nateglinide [Starlix] |
| Alpha glucosidase Inhibitor | miglitol [Glyset]; acarbose [Precose] |
| Thiazolidinedione | rosiglitazone [Avandia]; pioglitazone [ACTOS]; troglitazone [Rezulin] |
| Sodium-glucose cotransporter-2 Inhibitor | dapagliflozin [Farxiga]; canagliflozin [Invokana or Sulisent]; empagliflozin [Jardiance] |
| Dipeptidyl peptidase-4 Inhibitors | sitagliptin [Januvia]; saxagliptin [Onglyza]; linagliptin [Tradjenta] |
| Glucose-dependent insulintropic peptide-1 | exenatide [Byetta]; liraglutide [Victoza] |
| Insulin | regular insulin [Humulin R, Novolin R]; insulin lispro [Humalog]; insulin aspart [Novolog]; insulin glulisine [Apidra]; prompt insulin zinc [Semilente]; isophane insulin, neutral protamine hagedorn [NPH] [Humulin N, Novolin N]; insulin zinc [Lente]; extended insulin zinc insulin [Ultralente]; insulin glargine [Lantus]; insulin detemir [Levemir] |

^1^Evidence for specific prescriptions were identified using a master list of RxNorm and CUI codes for each of the medications listed in Table 2. Specific RxNorm and common unique identifier codes available upon request

**Table 3.** List of LOINC codes to identify HbA1c and HCG-B laboratory test values for the identification of patients with Type 2 diabetes and exclusion of patients with gestational diabetes

| Classification | Code list |  |
| --- | --- | --- |
| Hemoglobin A1c test | LOINC | 4548-4, 41995-2, 55454-3, 71875-9, 549-2, 17856-6, 59261-8, 62388-4, 17855-8 |
| Beta human chorionic gonadotropin (HCG) test | LOINC | 14041-8, 19174-2,19175-9, 19178-3, 19179-1, 19180-9, 20415-6, 2110-5, 2111-3,2112-1, 2113-9, 2114-7, 2115-4, 21198-7, 23841-0, 25373-2,29154-2, 32122-4,43799-6, 43800-2, 44003-2, 45293-8, 47024-5, 47601-0, 55866-8,55867-6, 55868-4, 55869-2, 56497-1 |

### Medication Covariates

| Medication class | Generic names [proprietary names] |
| --- | --- |
| Antithrombotic Therapy |  |
| Pharmacological Contraindications For Antithrombotic Therapy |  |
| Statin Grouper |  |
| Direct Thrombin Inhibitor |  |
| Glycoprotein IIb/IIIa Inhibitors |  |
| Injectable Factor Xa Inhibitor for VTE Prophylaxis |  |
| Low Dose Unfractionated Heparin for VTE Prophylaxis |  |
| Low Molecular Weight Heparin for VTE Prophylaxis |  |
| Oral Factor Xa Inhibitor for VTE Prophylaxis or VTE Treatment |  |
| Rivaroxaban for VTE Prophylaxis |  |
| Unfractionated Heparin |  |
| Warfarin |  |
| Dementia Medications |  |
| Antidepressant Medication |  |
| ACE Inhibitor or ARB or ARN |  |
| ADHD Medications |  |
| Substance Use Disorder Long Acting Medication |  |
| Substance Use Disorder Short Acting Medication |  |
| Tobacco Use Cessation Pharmacotherapy |  |
| Beta Blocker Therapy for LVSD |  |
| Beta Blocker Therapy |  |
| Antibiotic Medications for Pharyngitis |  |
| Contraceptive Medications |  |
| Isotretinoin |  |
| Anti Infectives, other |  |
| Anticholinergics, anti Parkinson agents |  |
| Anticholinergics, first generation antihistamines |  |
| Antipsychotic |  |
| Antispasmodics |  |
| Antithrombotic |  |
| Benzodiazepine |  |
| Cardiovascular, alpha agonists, central |  |
| Cardiovascular, other |  |
| Central nervous system, antidepressants |  |
| Central nervous system, barbiturates |  |
| Central nervous system, other |  |
| Central nervous system, vasodilators |  |
| Digoxin |  |
| Doxepin |  |
| Endocrine system, estrogens with or without progestins |  |
| Endocrine system, other |  |
| Endocrine system, sulfonylureas, long duration |  |
| Nonbenzodiazepine hypnotics |  |
| Pain medications, other |  |
| Pain medications, skeletal muscle relaxants |  |
| Reserpine |  |
| Pharmacologic Therapy for Hypertension |  |
| Aromatase Inhibitors |  |
| Glucocorticoids (oral only) |  |
| Adolescent Depression Medications |  |
| Adult Depression Medications |  |
| High Intensity Statin Therapy |  |
| Low Intensity Statin Therapy |  |
| Moderate Intensity Statin Therapy |  |
| Schedule II & III Opioid Medications |  |
| Schedule IV Benzodiazepines |  |
| Androgen deprivation therapy for Urology Care |  |
| BCG Bacillus Calmette Guerin for Urology Care |  |
| Chemotherapy for Advanced Cancer |  |
| Immunosuppressive Drugs for Urology Care |  |
| Medications for Above Normal BMI |  |
| Medications for Below Normal BMI |  |
| Anticoagulant Therapy |  |
| Thrombolytic (t-PA) Therapy |  |
| Hypoglycemics Severe Hypoglycemia |  |
| Hypoglycemics Treatment Medications |  |
| Anticoagulant Medications, Oral |  |
| Fibrinolytic Therapy |  |
| Total Parenteral Nutrition |  |

Medications will be included as clinical factor variables. These are the groups of medications that will be included for Propensity Scores and weights for Cox Proportional Hazards models, as these are groups of medications known to be associated with weight gain or weight loss (Singh).

New Section

- Interventions and Exoosures – cite design figure
